# The impact of tuberculosis and its treatment on the lung and gut microbiota: A global systematic review, meta-analysis, and amplicon-based metagenomic meta-analysis

**DOI:** 10.1101/2025.08.25.25334361

**Authors:** Monica Mbabazi, David Patrick Kateete, Faith Nakazzi, Joanitah Nabwire Wandera, Naume Mutesi, Moses Ocan, Irene Andia Biraro, Andrew Abaasa, William Evan Johnson, Bryan Wee, Adrian Muwonge

**Author notes:** Corresponding authors (DPK); (AM); (MM). First authorship (Monica Mbabazi).

## Abstract

**Background:** Tuberculosis (TB) remains the leading cause of bacterial disease-related deaths worldwide. Historically, the Koch’s single-causative-agent model has shaped diagnostics, treatment, and prevention. However, metagenomic studies have unveiled the presence of a lung microbiome, which is disrupted by TB and its orally administered treatment, with downstream effects on the gut microbiome. These changes may hold diagnostic, prognostic, and control potential once better understood. Here, we systematically analyze 38 studies with 3,394 individuals with TB and health controls to assess global insights on the impact of TB and its treatment on lung and gut microbiome diversity, structure, and composition. A meta-analysis with 24 studies estimates this effect size, while a patient-level amplicon metagenomic meta-analysis with 1617 individuals with 1.3 billion reads validates these associations. This study followed PRISMA guidelines and a pre-registered PROSPERO protocol (CRD42022329763).

**Results:** The systematic review reveals no global consensus on TB’s impact on the lung microbial diversity, though most studies report reduced diversity. However, we estimate a 0.14–0.41 overall reduction in lung and gut diversity. Patient-level lung diversity analysis showed no significant differences overall (Shannon index), though TB was associated with reduced diversity in China, but not in South Africa. In contrast, in the gut TB was associated with higher diversity in most countries. The TB diagnostic value of the microbiome remains uncertain, as disease status accounts for only 0.8–9% of lung and 1.8–9% of gut microbiota variation. However, lung depletion of *Prevotella, Neisseria, Veillonella, Haemophilus, Fusobacterium, Pseudomonas, Streptococcus, Porphyromonas*, and *Treponema*, along with gut depletion of *Prevotella, Ruminococcus, Faecalibacterium, Clostridium, Roseburia, Rothia, Eubacterium, and Escherichia*. TB treatment is associated with a reduction in diversity of both lung and gut.

**Conclusion:** TB is generally linked to reduced microbial diversity in the lung, but not gut. In contrast, treatment consistently reduces diversity in both, depleting key core genera. These findings underscore the exploitable potential of the gut–lung axis to improve TB diagnostics and prognosis.

## Background

With an estimated 1.25 million deaths in 2023, tuberculosis (TB) remains the world’s leading infectious cause of death (1). As of June 2024, the global fund to fight AIDS, TB and malaria has invested U.S. $9.9 billion in programs to prevent and treat TB, accounting for 76% of all international financing for TB (2). Additionally, the U.S. government increased its funding for global TB efforts from $242 million in 2015 to $406 million in 2024 (3). Yet, despite these substantial investments, TB continues to pose a major global health challenge. Historically, our understanding of TB remains rooted in Koch’s postulates of disease causation, identifying *Mycobacterium tuberculosis* (*Mtb*) as the predominant cause of TB in humans (4). However, advances in metagenomic sequencing have highlighted the microbiome’s role in health, disease pathogenesis, treatment outcomes, and sequelae (5). Indeed, similar patterns are noted for non-infectious diseases (6,7), supporting a notion that microbiomes retain signatures of disease more so for chronic infections like TB (8,9). Such signature represents diagnosis and prognosis clinical value. Microbiome studies offer a paradigm shift in understanding TB but are complex to interpret due to the breadth of host-pathogen-microbiota interactions (10). For example, the lung microbiota is dynamic and shaped by immune responses, upper respiratory tract migration (11,12), and clearance mechanisms, i.e. coughing, movement of respiratory cilia, pulmonary macrophages, alveolar surfactant-mediated bacterial inhibition (13) and microbial migration (14). Disruptions in any of these processes can alter microbial composition (15,16), reducing resistance to colonization by non-resident microbes. Indeed, health is often linked to a diverse microbiota, while dysbiosis, defined as microbiota imbalance and loss of diversity, is associated with disease (17,18). In the lungs, current evidence suggests this is shaped by immune reorganization, whereas in the gut, it results from prolonged TB medication. This therefore raises key questions:

i. What are the key compositional differences between the lung and gut microbiomes of TB patients and healthy individuals? (ii) Are there consistent microbial biomarkers across different geographic regions? (iii) How does anti-TB treatment impact microbiota composition? Answering these questions will enhance our understanding of the microbiota’s role in active TB onset, progression, and treatment response. Importantly, this knowledge can inform the development and use of interventions such as prebiotics and probiotics. The challenge lies in disentangling these overlapping microbial signatures when multiple factors affect an individual simultaneously.

Here, we use a mixed-methods synthesis of literature to examine key compositional differences between TB cases and healthy controls, and assess whether these signatures are consistent across geographic regions. We also examine how anti-TB treatment alters the microbiota, aiming to identify key microbial taxa associated with disease and treatment response.

To date, studies investigating the lung and gut microbiota in TB patients have been conducted across a wide range of geographic regions. **Figure 1** illustrates the global distribution of the studies included in this review, highlighting the global relevance of microbiota research in TB. This wide geographic spread is important for assessing both shared and region-specific microbial signatures and their implications for disease understanding and intervention strategies.

**Figure. 1:**
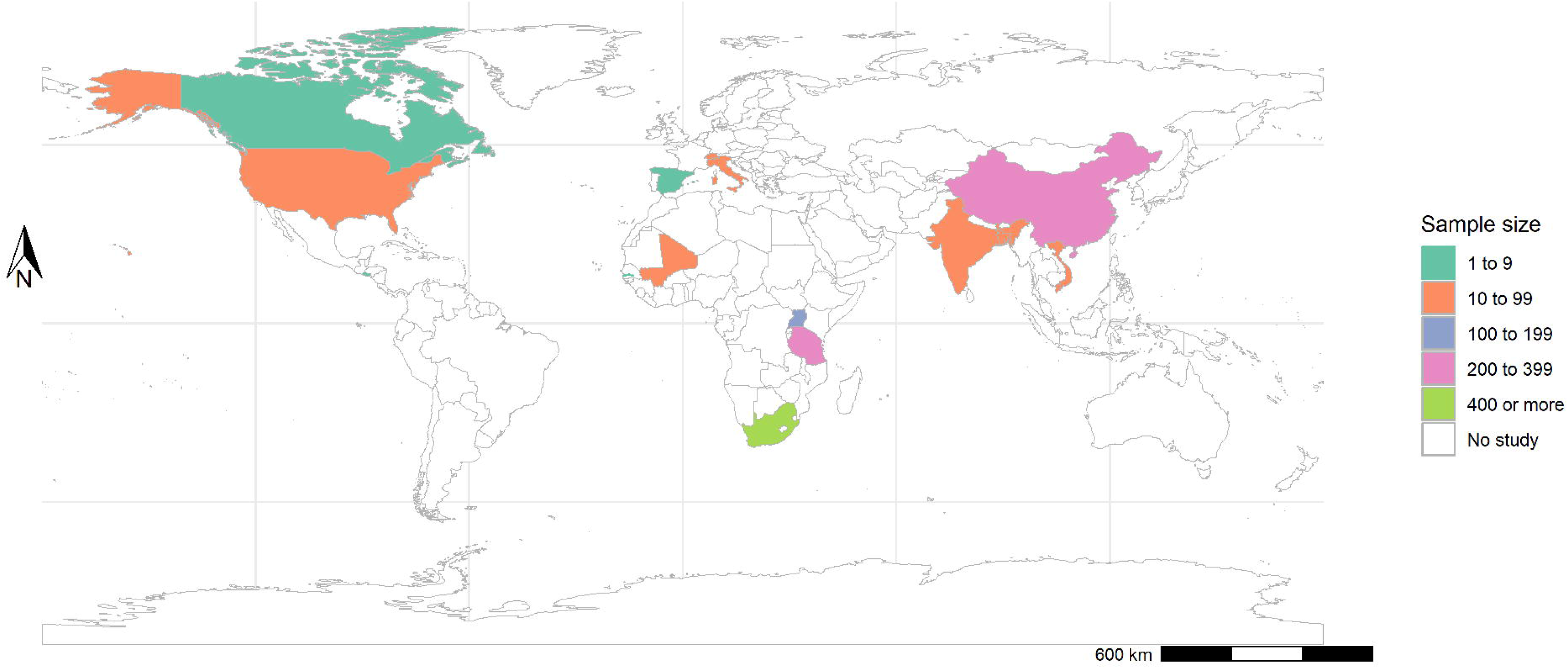
Geographic distribution and sample sizes of studies included in the systematic review and meta-analysis. This map illustrates the countries from which microbiome data were extracted in the review, categorized by total sample size. Colour coding indicates the number of participants included per country, ranging from fewer than 10 to more than 400. Countries shaded in white had no eligible studies meeting the inclusion criteria. This global distribution highlights the geographical diversity of available data and supports regional comparisons in lung and gut microbiota associated with tuberculosis. Asia contributed the largest number of samples, followed by Africa and the Americas, with Europe contributing the fewest.

This raises several key questions:

1). What are the key compositional differences between the lung and gut microbiomes of TB patients and healthy individuals?

2). Are there consistent microbial biomarkers across different geographic regions?

3). How does anti-TB treatment impact microbiota composition?

Answering these questions will enhance our understanding of the microbiota’s role in TB onset, progression, and treatment response. Importantly, this knowledge can inform the development and use of targeted microbiota-based interventions such as prebiotics and probiotics. The challenge lies in disentangling overlapping microbial signatures in individuals exposed to multiple interacting factors. To address this, we employed a mixed-methods synthesis of the literature, integrating three complementary analytical approaches to examine shifts in microbiota diversity, structure, and composition: (a) a systematic review to assess consensus on microbial differences between healthy controls, TB cases, and treatment status; (b) a meta-analysis to quantify effect sizes; and (c) an amplicon metagenomic meta-analysis at the patient level to validate and visualize patterns. **Figure 2** outlines the conceptual framework guiding this analysis.

**Figure. 2:**
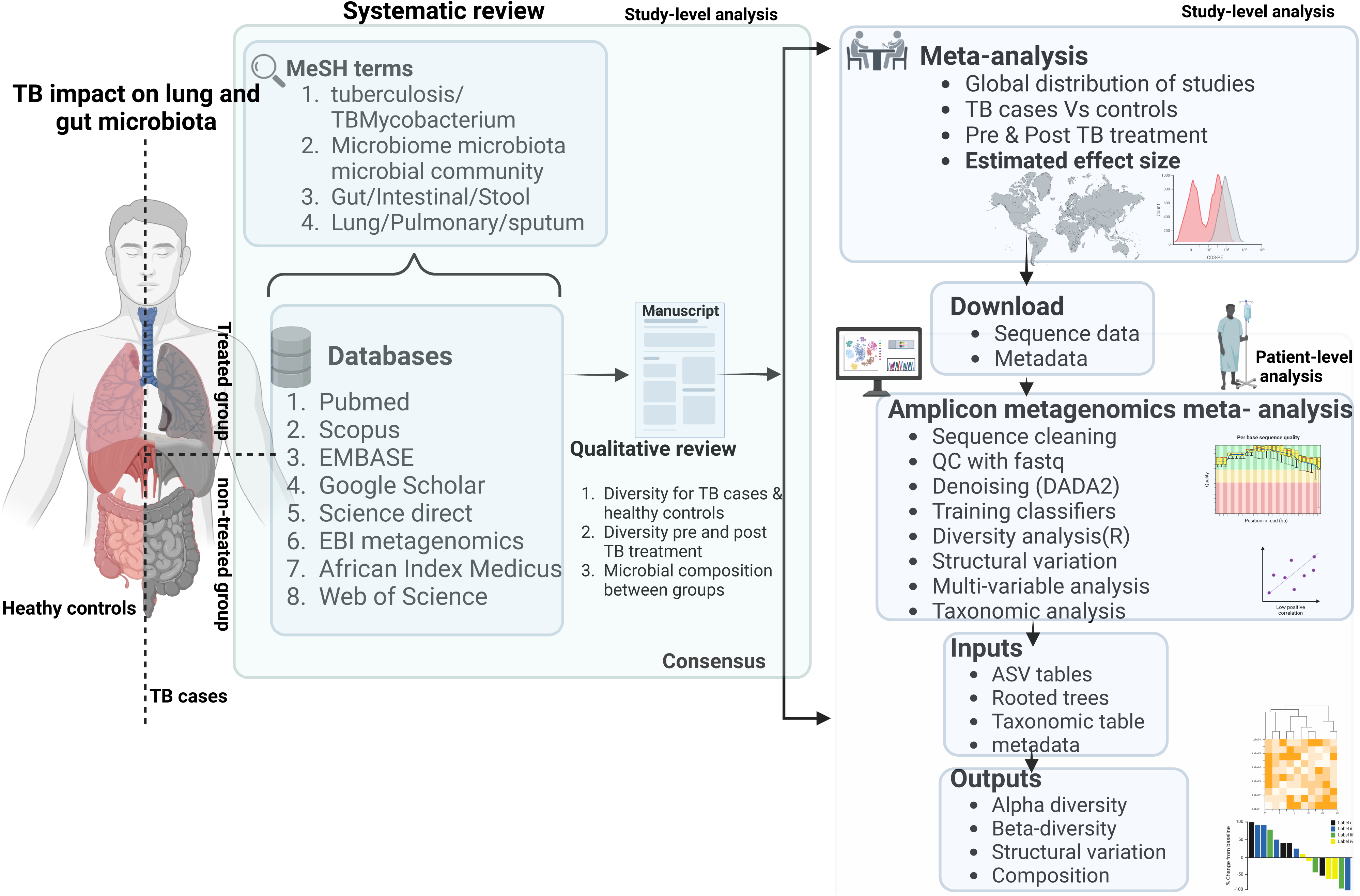
This conceptual flow diagram summarizes the three main methodological components of the study: (1) **Systematic review** to identify trends in the microbiome and TB research, (2) **Meta-analysis** of reported alpha diversity measures to quantify microbiome shifts in TB cases and treatment groups, and (3) **Amplicon-metagenomic meta-analysis (AMMA)** to analyze raw sequencing data from publicly available datasets. Each stage outlines the data sources and key analytical steps performed.

## Methods

### Study design and literature search

This study followed PRISMA guidelines and a pre-registered PROSPERO protocol (CRD42022329763). It integrated a systematic review to identify trends, a meta-analysis to estimate effect sizes, and a patient-level amplicon metagenomic meta-analysis (AMMA) for validation. A comprehensive search of peer-reviewed studies on gut and lung microbiomes in TB patients and healthy controls, including treatment effects, was conducted up to 2023. Studies without healthy controls were included if they assessed treatment impacts. Eligible studies underwent rigorous screening for narrative synthesis and quantitative analysis.

### Search terms and strategy

A systematic search was conducted across multiple electronic databases, including PubMed, Scopus, EMBASE, Google Scholar, ScienceDirect, EBI Metagenomics, AIM (African Index Medicus), and Web of Science, for studies published in English. The search strategy combined Medical Subject Headings (MeSH) terms and free-text keywords, and was supplemented by hand-searching reference lists, related articles, and relevant gray literature. The complete list of MeSH terms and keyword combinations used is provided in **Supplementary Table S1**. The search was independently performed by three reviewers (MM, FN, and JN), and any discrepancies were resolved through discussion with a fourth expert reviewer (DPK).

### Inclusion and exclusion criteria

The study employed three analytical approaches: systematic review, meta-analysis, and AMMA, each with distinct inclusion criteria. The systematic review included all study designs; observational and interventional, focusing on lung and gut microbiomes. Various biological samples, such as BAL, nasal/oropharyngeal swabs, sputum, and stool, were analyzed. Studies included TB patients, with or without healthy controls, and those examining treatment effects. The meta-analysis included only systematic review studies reporting the mean Shannon index for TB cases and controls, as well as treated and untreated groups. Effect sizes were computed and visualized as forest plots.

For the AMMA, only studies from the systematic review that provided raw FASTQ files of 16S rRNA sequences and metadata were included, regardless of the targeted gene region. Exclusion criteria varied by approach. The systematic review excluded animal studies, case reports, in vitro studies, abstracts, protocols, reviews, letters, inaccessible full texts, and studies on non-lung/gut microbiota, including extra-pulmonary TB. The meta-analysis excluded studies lacking TB cases and controls, those without Shannon index values, or those not distinguishing treated from untreated TB cases. The AMMA excluded shotgun sequencing studies and amplicon studies without raw reads, considering only paired-end reads.

### Eligibility screen

1. Articles from the literature search were imported into Mendeley(19) , and duplicates were removed by three independent reviewers (MM, FN, JN).
2. Title & Abstract Screening: Three reviewers independently screened titles and abstracts for eligibility. Disagreements were resolved by a fourth reviewer (DPK).
3. Full-Text Screening: Full texts of selected studies were assessed for eligibility, with unclear cases reviewed by the fourth reviewer (DPK).

### Data extraction

After selecting eligible studies, data was extracted and entered in a structured spreadsheet, categorized by key variables, including study details, methodology, TB treatment status, sequencing platform etc. (Supplementary_Data.xlsx). Study quality was assessed for validity, transferability, size, and precision.

### Data synthesis and analysis

**Systematic review:** Tables S2 and S3 summarize the dataset by publication year, location, sample size, sequencing approach, and microbiome differences in TB and its treatment. Table S2 covers lung microbiome studies, while Table S3 the gut microbiome studies.

**Meta-analysis:** Shannon diversity values were used to compute effect sizes, evaluating TB’s impact (cases vs. controls) and treatment effects (treated vs. untreated) on microbiome diversity. Separate meta-analyses for gut and lung microbiomes were visualized as forest plots.

**AMMA:** Reads from 11 studies meeting inclusion criteria were downloaded, quality-controlled, and linked to standardized metadata. Raw sequences were processed using QIIME 2 (19,20), with quality control ensuring Phred scores ≥20 and read trimming to 200 base pairs. DADA2 was used to obtain representative amplicon sequence variants (ASVs) from 1,885 samples, analysed in batches of 11 before merging for diversity, phylogenetic, and taxonomic assessments.

Taxonomic classification was harmonised across variable regions using the full-length 16S rRNA gene-trained classifier. The ASV table, taxonomic table, rooted tree, and metadata were merged using the phyloseq package (21) in R 3.5.1. Contaminants, eukaryotic reads, and samples with <500 reads were removed. Alpha and beta diversity were analysed using the Microbiome package in R, visualized via boxplots and PCA plots in ggplot2. Microbiota structure associations were assessed using PERMANOVA, while DESeq2 in Bioconductor(22). identified differentially abundant taxa between TB cases and controls, as well as treated versus untreated groups.

## Results

### A summary of the study eligibility screening

A literature search yielded 987 records, with 652 excluded based on species, age, or language criteria. After removing nine duplicates, the remaining records were screened using PRISMA guidelines, which excluded 182 based on titles and abstracts. Of 144 full-text articles assessed, 106 were excluded using the Cochrane PICOS (Population Intervention Comparison Outcomes and Study design) model. Following independent reviewing, 38 studies were included in the systematic review. Further screening identified 31 studies for meta-analysis and 11 for metagenomic meta-analysis, as detailed in the PRISMA flowchart (**Figure 3**).

**Fig. 3:**
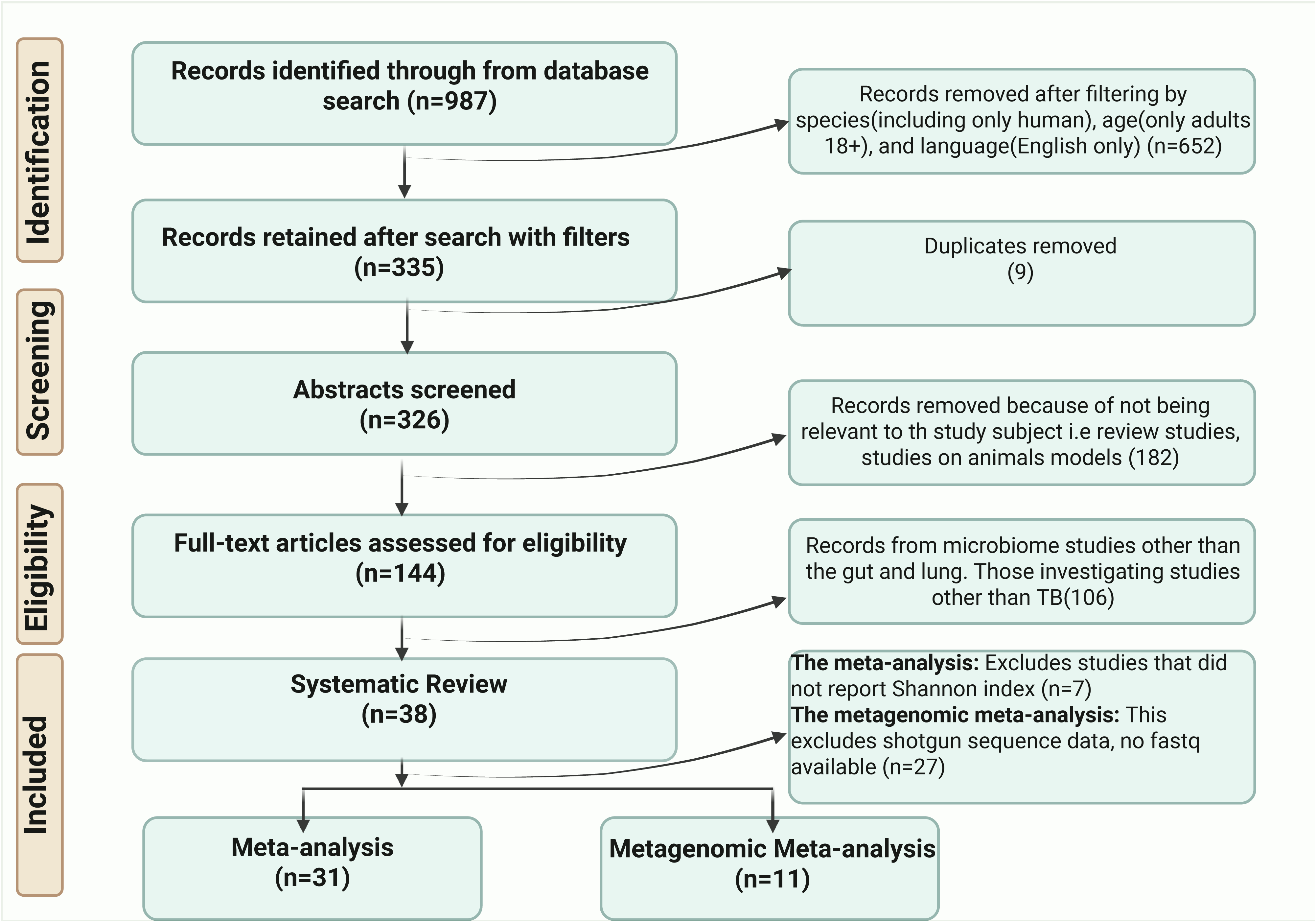
PRISMA flow diagram showing the selection criteria. This diagram illustrates the stepwise process of identifying, screening, and selecting studies that met the eligibility criteria. It also highlights the number of studies retained for the systematic review, meta-analysis, and metagenomic meta-analysis after applying the inclusion criteria.

### Characteristics of microbiota studies of tuberculosis

A total of 3,394 participants from studies published up to 2023 were included. Twenty studies (1,903 TB patients, 426 controls) examined lung microbiota, and 17 (630 TB patients, 435 controls) focused on gut microbiota. One South African study analyzed both. Most studies were cross-sectional, with sample sizes ranging from 12 to 384. Asia contributed the most samples, Europe the least. Sputum was the most common respiratory sample, while gut studies used stool. Only five studies used shotgun sequencing; most relied on amplicon sequencing, primarily targeting the V3-V4 region of the 16S rRNA gene (Table S 2&3).

To investigate patient level variability, we retrieved 1.3 billion raw reads (∼276 GB) from GenBank, covering 1,617 samples from 11 amplicon metagenomic studies for a patient-level meta-analysis of lung (5 studies, 1,067 participants) and gut microbiota (6 studies, 550 participants). Lung microbiota diversity was higher in Africa and Asia than in Europe and America (**Figure S1**).

### Lung and gut dysbiosis associated with tuberculosis

In the lung, TB cases exhibit enrichment of anaerobic genera (*Streptococcus, Haemophilus, Oribacterium*, and *Veillonella*) and depletion of aerobic genera (*Neisseria, Micrococcus, Nocardia*, and *Moraxella*). Healthy controls on the other hand exhibit enrichment of *Prevotella, Treponema, Leptotrichia, Lactobacillus*, and *Actinobacillus* (Table S1&2) as reported. AMMA confirms the overall depletion of *Prevotella, Neisseria, Porphyromonas*, and *Treponema* in TB cases (Fig 6). This result however suggests that *Veillonella and Haemophilus* are depleted in TB cases. TB appear to alter gut microbiota composition, with cases showing depletion of *Roseburia*, *Eubacterium*, *Faecalibacterium*, *Escherichia*, *Bifidobacterium*, and *Clostridium*. AMMA reveals a general depletion of *Romboutsia*, *Sutterella*, *Campylobacter*, *Agathobacter*, and *Prevotella*, which are enriched in healthy controls.

### Global consistency of TB associated microbial composition and diversity

Pulmonary TB is associated with a depletion of *Prevotella and Treponema*, while these genera are enriched in healthy controls. Country-specific microbial signature observed: in China, where TB patients exhibit additional depletion of *Neisseria*, *Campylobacter, Fusobacterium, Leptotrichia, and Moraxella*; in Bangladesh, healthy controls exhibit enrichment of *Alloprevotella, Oribacterium, Burkholderia, Gemella, and Peptostreptococcus* (Fig. 6). In contrast, no significant microbial diversity or compositional differences were observed in South Africa.

Gut microbiota profiles also vary by country. In China, TB cases exhibit enrichment of *Escherichia-Shigella, Subdoligranulum, and Faecalibacterium*, while controls it is *Bifidobacterium* which is enriched. In the U.S., TB cases exhibit enrichment of *Bacteroides, Dialister* and *Succinivibrio*, whereas healthy controls are enriched with *Prevotella, Bacteroides spp*., and *Sutterella*. In South Africa, TB cases are enriched with *Gemella, Streptococcus, and Neisseria*, and here too *Prevotella* predominates in controls.

The above compositional shifts are generally exhibited as reductions in the Shannon index among TB patients as revealed by the meta-analysis of 38 studies (lung: 20, gut: 20, both: 1). Indeed, most studies support this trend, though three from China reported higher lung microbiota diversity in TB cases, while two from South Africa and the U.S. found no significant difference (Table S1&S2). A similar diversity pattern is observed in gut microbiota, with some contradictory findings. Overall the meta-analysis of 10 lung studies shows a 0.14 reduction in the mean Shannon diversity index, while gut microbiota diversity is more affected, with a 0.41 reduction among TB cases (**Figure 4**).

**Fig. 4:**
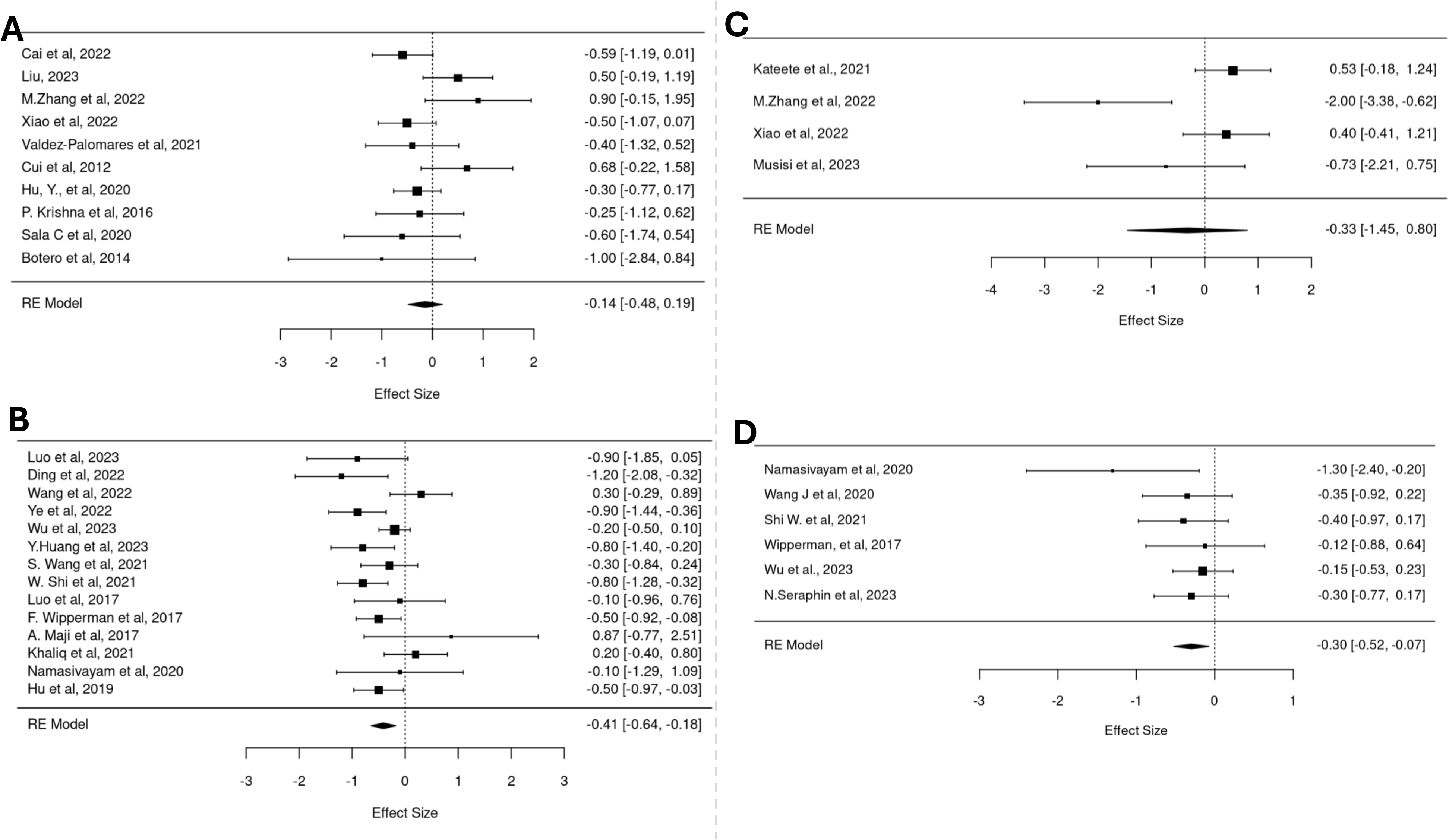
Forest plots summarize the impact of tuberculosis and its treatment on lung and gut microbial diversity. **(A)** TB is associated with a modest reduction in lung microbial diversity (effect size: –0.14; 10 studies). **(B)** TB is linked to a more pronounced reduction in gut microbial diversity (effect size: –0.41; 14 studies). **(C)** Anti-TB treatment further reduces lung microbial diversity (effect size: –0.33; 4 studies). **(D)** Anti-TB treatment is also associated with decreased gut microbial diversity (effect size: –0.30; 6 studies). All estimates are based on meta-analyses using random-effects models.

### Patient-level differences linked to tuberculosis

While no significant difference in Shannon diversity was found between TB cases and controls **(Figure 5**), TB cases showed greater variability (Fig 5A). In China, TB cases had lower Shannon diversity, whereas no differences were observed in South Africa and Bangladesh (Fig 5C). In contrast, TB significantly affected gut microbiota, with cases exhibiting higher Shannon diversity across China, South Africa, India, and the U.S., challenging previous reviews (Fig 5B &C). Faith’s phylogenetic and beta diversity analyses revealed that TB cases had lower lung but higher gut microbiota diversity. These differences are shown as distinct clustering by disease status using PCA, and PERMANOVA indicating that disease status explained 0.8–9% of lung and 1.8–9% of gut microbiota variation **(Figure 6**). However, geography and sequencing methods also contributed to the variation, with continent and 16S rRNA target regions accounting for 1.2% and 8.9% of lung microbiota differences, respectively (Table 1). This finding suggests that TB exerts opposing effects on lung and gut microbial communities.

**Figure 5:**
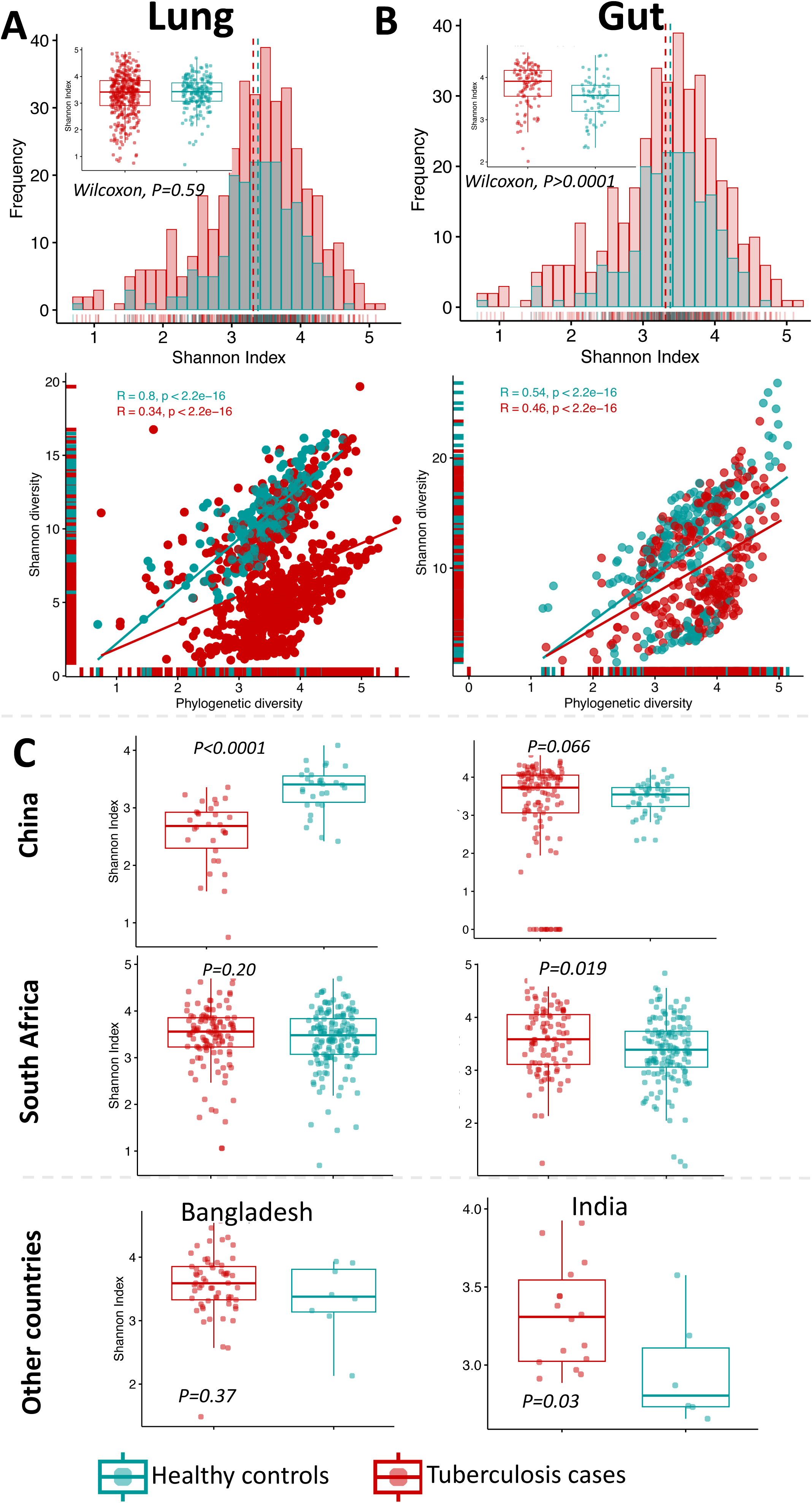
The divergent effects of tuberculosis on lung and gut microbiome diversity (Shannon index). While the lung microbiota diversity shows country-specific variations in TB cases compared to healthy controls, the gut microbiota consistently exhibits higher diversity in TB cases across all regions.

**Figure. 6:**
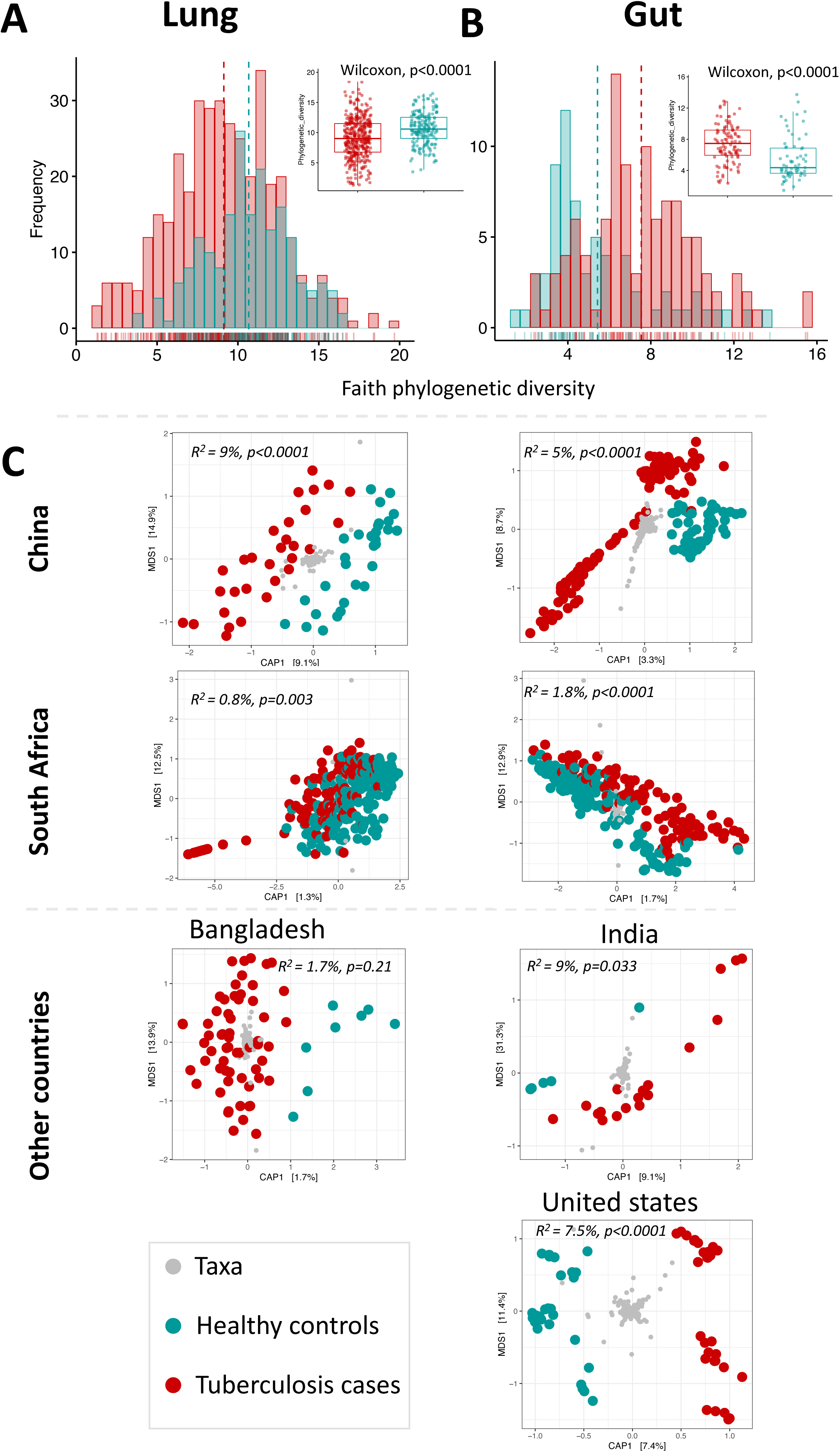
**A distinct TB signal in the lung and gut microbiomes revealed using Faith’s phylogenetic diversity and PCA analysis**. This divergence alluded to in Fig 5 is also reflected in lower and high Faith’s phylogenetic diversity of the lungs and gut of TB patients respectively compared to healthy controls. PCA plots demonstrate clustering by disease status, distinguishing TB cases from healthy controls. This distinct clustering suggests a significant impact of TB on both lung and gut microbiomes.

**Table 1:**
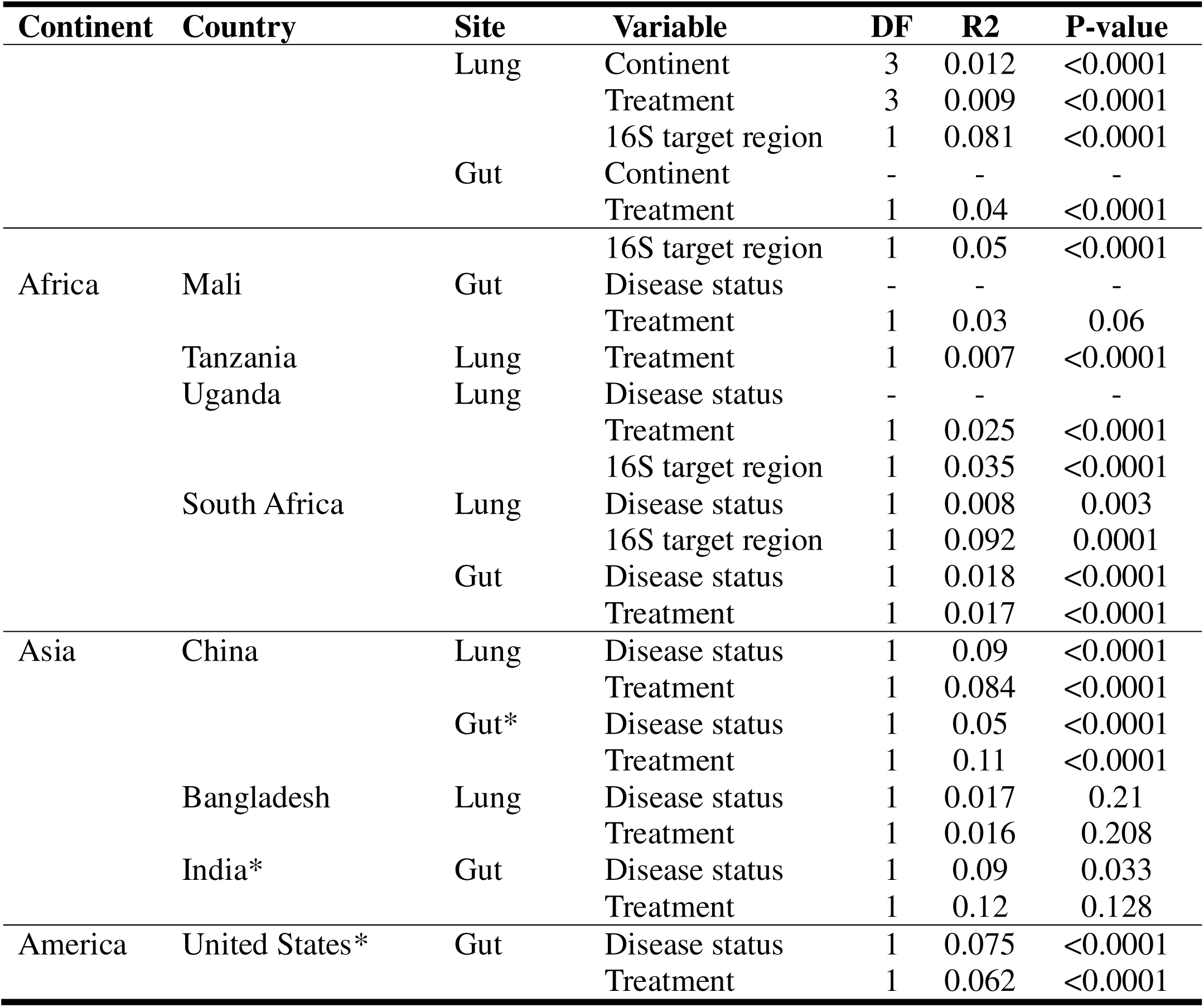
Assessing the variation explained by “place” at the continent and country levels, as well as the influence of microbiome anatomical sites, targeted sequencing variable region, and TB management using PERMANOVA.

### Lung and gut dysbiosis linked to anti-TB treatment

Anti-TB treatment are associated with significant alterations in lung microbiota, with treated patients exhibiting depletion of *Fusobacterium, Veillonella, Prevotella, Streptococcus, Gemella*, and *Leptotrichia*, which are enriched in untreated cases (**Figure 7**). Here too we observe country-specific effects which include enrichment of *Blautia* and *Bacteroides* in treated patients in China, while *Prevotella* becomes relatively in untreated groups. In Tanzania, untreated individuals exhibited enrichment of *Neisseria, Streptococcus, Gemella, Campylobacter*, and *Fretibacterium*. In Uganda, treatment enriches *Streptococcus spp*. and *Alloprevotella* while depleting *Haemophilus* and *Neisseria*. In the gut, there is a general enrichment of *Prevotella, Succinivibrio*, *Faecalibacterium*, *Dialister*, and *Alloprevotella* among the non-treated groups. In India, treated groups exhibit enrichment of *Clostridium*, *Christensenella*, *Fusobacterium*, and *Eubacterium*, while in Mali, treatment is associated with *Prevotella* and *Agathobacter* enrichment.

**Figure. 7:**
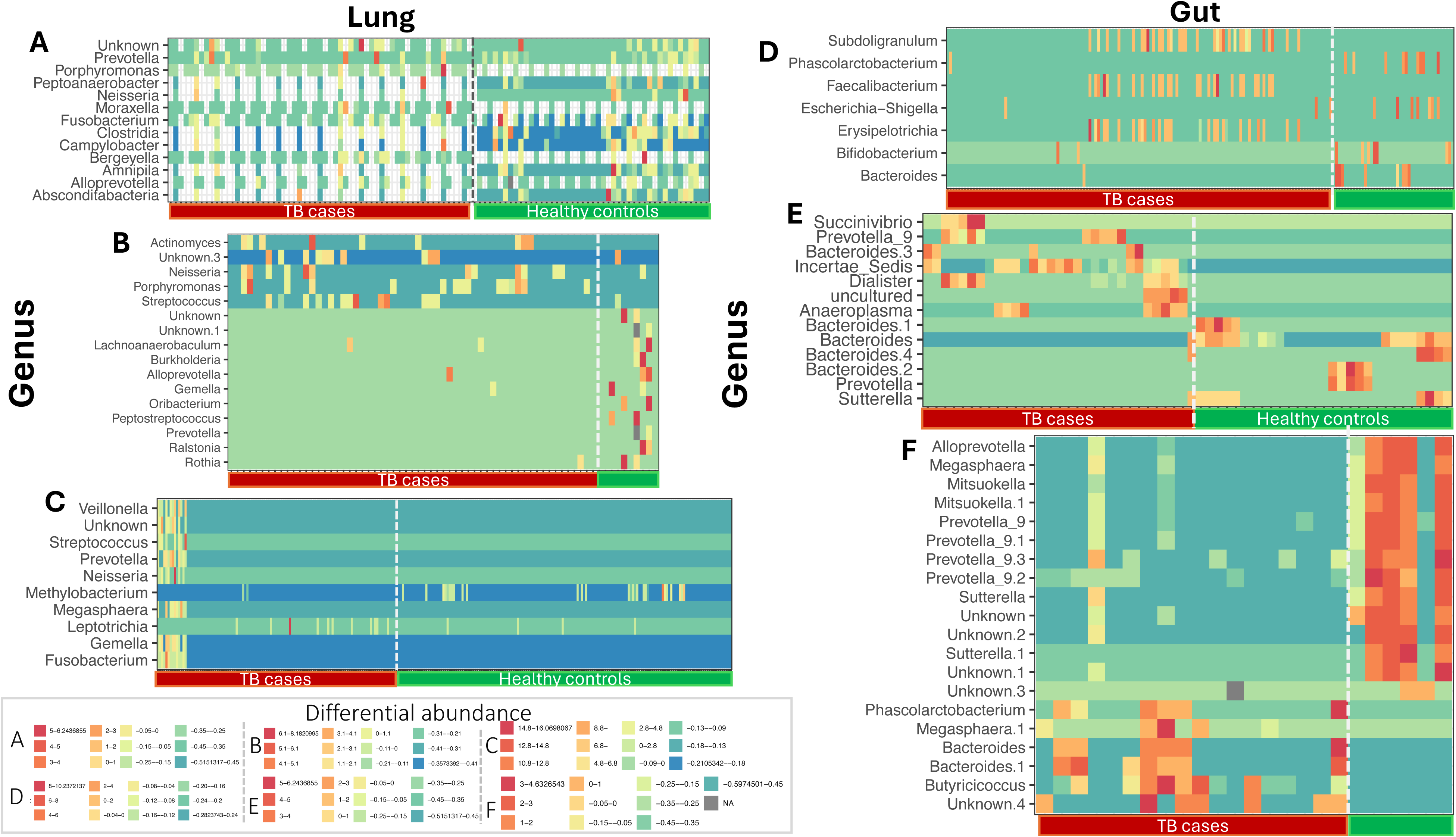
Heatmap depicting the differential abundance of microbial taxa in lung and gut samples. In lung microbiota, TB cases exhibit enrichment of anaerobic genera (e.g., Streptococcus, Haemophilus, Oribacterium, Veillonella) and depletion of aerobic genera (e.g., Neisseria, Micrococcus, Nocardia, Moraxella), while healthy controls are enriched with Prevotella, Treponema, Leptotrichia, Lactobacillus, and Actinobacillus. In gut microbiota, TB cases show significant compositional alterations compared to healthy controls, with distinct patterns observed across different regions and treatment statuses.

### Patient-level variations and anti-TB treatment

There is strong consensus in literature that anti-TB treatment reduces lung and gut microbiota diversity, and indeed the meta-analyses of four lung and six gut microbiota studies highlights this trend. Additionally, the patient level analysis of five studies including 874 participants with pulmonary TB from eleven countries across four continents, reveals a significant difference of phylogenetic composition between treated and untreated groups (**Figure 8**). At country level, this effect was evident in Tanzania but not in Uganda. A similar pattern was observed in gut microbiota (Figure 8).

**Figure. 8:**
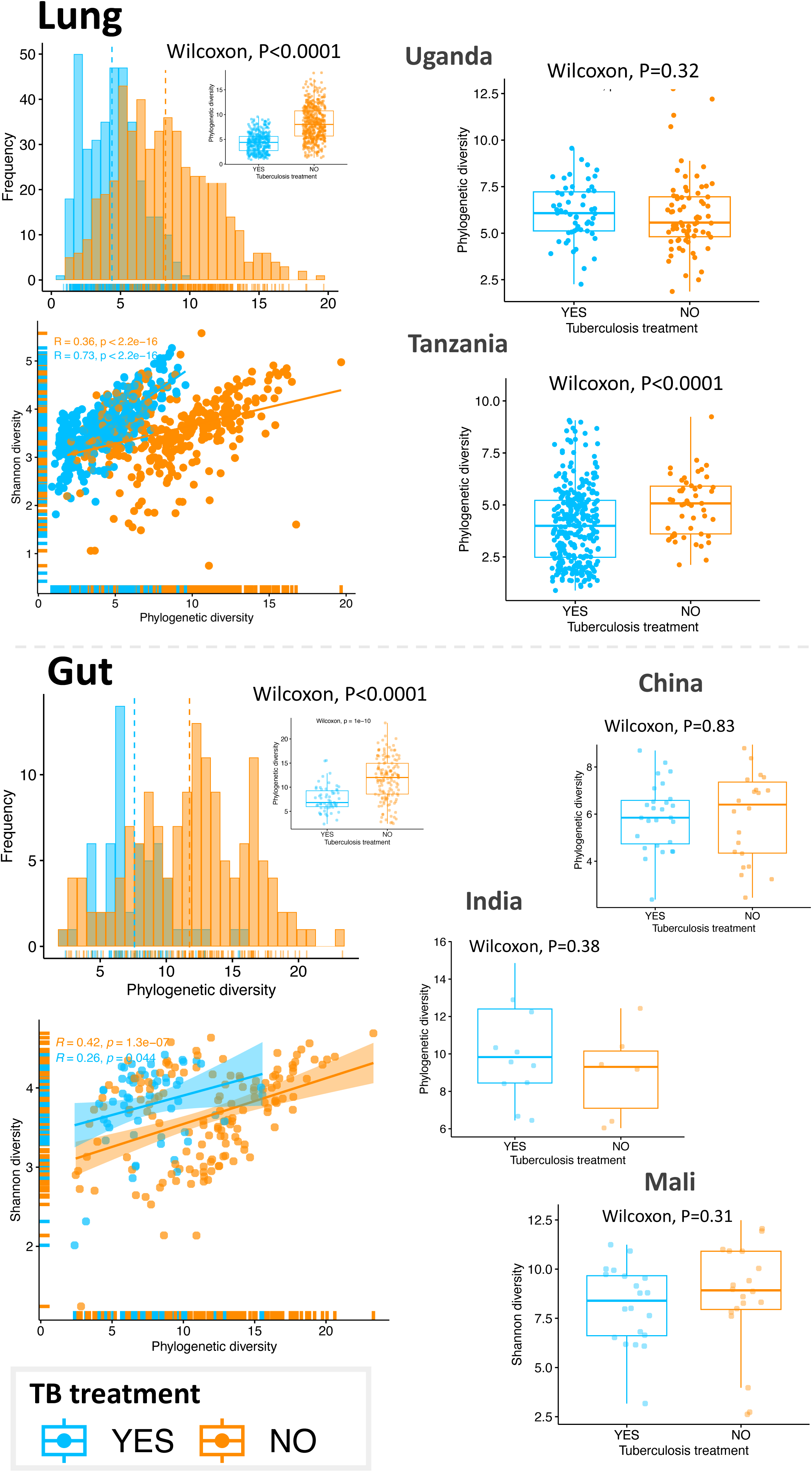
TB treatment is consistently associated with a lower lung and gut microbiota diversity when compared to untreated individuals. Overall. However, country-specific variations are observed: in lung microbiota, treated individuals in Uganda show higher diversity, while those in Tanzania exhibit lower diversity. For gut microbiota, higher diversity in treated individuals is observed only in India, whereas in China and Mali, untreated individuals display greater diversity.

## Discussion

Historically the lung was considered sterile but now, through metagenomics— it is known to host a low-biomass, low-diversity microbiome, connected to the gut via the gut-lung axis(22,23). In healthy individuals, microbial composition is reported to be maintained by physiological migration between the oropharynx, upper and lower respiratory tracts (14). However, the disease mechanisms underlying dysbiosis (shifts in microbiota composition) remain under active investigation as understanding lung-gut microbiota composition and diversity is key to leveraging microbial signatures for TB diagnostics and management. It is noteworthy that variability in observations is influenced by sequence targeting methodology.

Here we harmonized the analysis of variable 16S rRNA target region to reveal the following key findings: a) The association between TB and lung microbiota diversity is less consistent than for the gut, with most studies showing reduced diversity but some reporting no effect or the opposite. b) TB generally reduces lung diversity but increases gut diversity, c) Gut microbiota findings show consistency in trends, as we estimate a 0.41 and 0.30 reduction in diversity of lung and gut microbiota. d) Taxonomic composition is associated to TB, exhibiting depletion of resident genera previously associated with pro- and anti-inflammatory effects in the lung and gut, respectively.

### A reduction in lung microbial diversity may be associated with tuberculosis

The meta-analysis findings here suggest that TB impacts the lung microbial diversity, with an estimated reduction of 0.14 in the Shannon diversity index. However, the patient-level analysis revealed no significant difference between TB cases and health controls. It is likely that variability in 16S rRNA targets contribute to this this inconsistency although when contrasted with the Faith’s phylogenetic diversity (PD) there is a consistent reduction in diversity among TB cases both in national and international comparison. On the other hand, TB appears to have the opposite effect in the gut when assess using Faith’s PD. Some studies have argued that the reduction in lung microbial diversity may result from immune responses (12,24,25), where alveolar macrophages eliminate both Mtb and resident microbes (26), which is likely here captured as the impact size. We also note that TB cases show greater variability in diversity indices compared to healthy controls, suggesting more stable microbiota diversity in healthy individuals. We therefore argue that variability between cases i.e., beta diversity might be a more reliable measure of TB’s impact on lung microbiota. Indeed, when analyzed this diversity revealed a consistent disease-related signal globally and within countries. We estimate that TB explains 0.8–9% of lung microbiota variation, here the strength of association is greater at country level. Overall, we show that TB reduces the frequency of phylogenetically distant bacteria in the lung but increases it in the gut. We therefore posit that this divergence in response may represent exploitable biomarkers for TB management. Furthermore, the use of beta diversity indices might could enhance the diagnostic values of lung and microbiota.

Our patient-level meta-analysis shows TB-related lung microbiota shifts, marked by the depletion of core genera like *Prevotella, Neisseria, Veillonella, Haemophilus, Fusobacterium, Pseudomonas, Streptococcus, Porphyromonas*, and *Treponema* (27). This contrasts with reports of increased *Streptococcus, Prevotella, Veillonella*, and *Atopobium* in sarcoidal conditions, an example of which is TB (28), warranting further investigation. Since core lung microbiota modulates immune responses via IL-17 and T helper cells (12), this depletion may impact immunity and disease prognosis (29). TB-HIV co-infection and *Prevotella* enrichment predict mortality, while smoking depletes *Porphyromonas*, *Neisseria*, and *Gamella* (30), though our analysis does not account for such interactions.

### TB is associated with higher gut microbial diversity

While some studies report higher gut microbiota diversity in TB cases, most show the opposite, quantified as 0.41 reduction in the Shannon index. However, patient-level analysis of six studies (550 individuals) indicates greater gut diversity in TB cases, an observation supported by the multivariable regression-based analysis of evenness and phylogenetic richness. In this regard the Beta diversity suggests that TB explaining 1%–9% of gut microbiota variation across all countries, however refining estimates may require robust metadata such as malnutrition, alcohol use, and diabetes (31). Unlike lung shifts likely driven by *M. tuberculosis*, gut microbiota changes remain unclear, but other studies show it involves alteration in core genera composition like *Prevotella*, *Ruminococcus*, *Faecalibacterium*, *Clostridium*, *Roseburia*, *Rothia*, *Eubacterium*, *Escherichia*, and *Subdoligranulum* (*32–34*). Notably, here TB cases show *Faecalibacterium* and *Roseburia* depletion which have been linked to short chain fatty acid production associated with IL-10-mediated immunity(ref). In China, TB is linked to *Escherichia* and *Subdoligranulum* enrichment in the gut, which is associated with protracted dysbiosis and anti-inflammatory responses (35,36).

### TB treatment is associated with reduced lung and gut microbial diversity

Antibiotic use alters the composition and function of lung microbiota (37), but the duration of these changes remains uncertain. TB treatment regimens six (180 doses) vs. nine (780 dose) months) impact lung microbiota (31), consistently reducing diversity by depleting core genera like *Fusobacterium, Veillonella, Prevotella, Streptococcus, Gemella,* and *Leptotrichia*. This diversity reduction likely arises from impaired pulmonary clearance (37) or broad-spectrum antibiotics like Rifampicin which can eliminate core microbiota. Such compositional shifts likely affect lung immunity (38), for example; *Prevotella* depletion is linked to increased Th17-mediated inflammation and reinfection risk, while *Gemella* loss may impair lesion absorption by altering interferon-gamma modulation (39). Specifically, *Prevotella* depletion is linked to increased Th17-cell-mediated inflammation and increased reinfection risk (40). Furthermore, the combined loss of *Veillonella*, *Prevotella*, and *Streptococcus* is reported to disrupt anti-inflammatory macrophage differentiation, affecting anabolic pathways (41). Country-specific variations exist; in Uganda, TB treatment enriches *Streptococcus* while depleting *Haemophilus*. Although lung microbiota reduction in TB is less pronounced than in the gut, its immunological impact may influence inflammation, disease progression, and reinfection risk, with variability across settings.

TB treatment on the other hand is also shown to consistently reduce gut microbial diversity across spatial and temporal scales. Early studies on first-line TB drugs (*isoniazid, pyrazinamide, ethambutol*) reported gut dysbiosis without a clear trend, but subsequent studies have linked the prolonged use to lower richness and Shannon diversity (42). Our patient level analysis reveals a similar trend with Faith’s Phylogenetic Diversity. Here we show that treatment depletes *Prevotella, Bacteroides, Faecalibacterium, Succinivibrio*, and *Clostridium*. *Prevotella* depletion is reported to be a correlate for immune markers such CD4+ lymphocyte counts (43). On the other hand, *Bacteroides* abundance is associated with disease progression but also plays a key role in microbial nutrient mobilization and immune modulation (43). Geographic gut microbial variability also shapes this relation, for example in India and Haiti (44), TB patients show *Clostridium* and *Erysipelatoclostridium* enrichment, like findings from mouse models where *Clostridium* declines early and late in treatment (45). Over all such shifts can disrupt carbohydrate metabolism and immune regulation along the gut-lung axis, which are reported to increase post-treatment reinfection risk. Indeed, reinfection has been linked to depleted T-cell epitopes from non-tuberculous mycobacteria, with our results showing a significant reduction in Mycobacteria among treated individuals. These patterns suggest TB may have pro-inflammatory effects in the lung but anti-inflammatory effects in the gut.

### A paucity of evidence on effects of HIV/TB co-infection on the microbiome

Despite antiretroviral scale-up reducing HIV infections and mortality, HIV-associated TB remains a major challenge, especially in low-income countries. TB is the leading opportunistic infection and cause of death among people with HIV, yet microbiome studies on this co-infection are limited. The lack of clear differentiation between TB cases and healthy controls in South Africa may be linked to HIV-related confounding. Further research in high TB-HIV burden settings is needed to clarify microbiome interactions with TB progression and immunity.

### Clinical, practical relevance of findings & future research priorities

***Probiotics in TB Management***: The observed 0.14 and 0.41 reductions in lung and gut diversity highlight the potential of probiotics as pre- and post-treatment strategies to restore microbial balance (Figure 7).

***Microbiota as a Diagnostic & Prognostic Tool***: Amplification of this signal could be achieved by applying machine learning on Lung and gut microbiota to enhance TB diagnostics, disease progression monitoring, and treatment response assessment.

***Immune Modulation***: Some microbial landscapes identified are linked to metabolites like short-chain fatty acids and peptidoglycans, which activate toll-like receptors (TLRs) and influence immune responses (42). Future studies should explore these pathways as potential complementary therapies for TB targeting pro and anti-inflammatory cascades (46).

***Need for Longitudinal Studies***: Understanding microbiota dynamics, particularly treatment-induced shifts, is crucial for patient welfare.

***Advancing Clinical Metagenomics***: Investigating microbiota-driven treatment outcomes, Mtb lineage variations, and geographic influences could drive innovations in both population-level and personalized TB management.

These conclusions are visually summarized in **Figure 9**, which consolidates our key findings across lung and gut microbiota in TB patients, detailing changes in diversity, microbial structural variation, and taxonomic profiles based on systematic review, meta-analysis, and amplicon metagenomic analysis.

**Figure 9:**
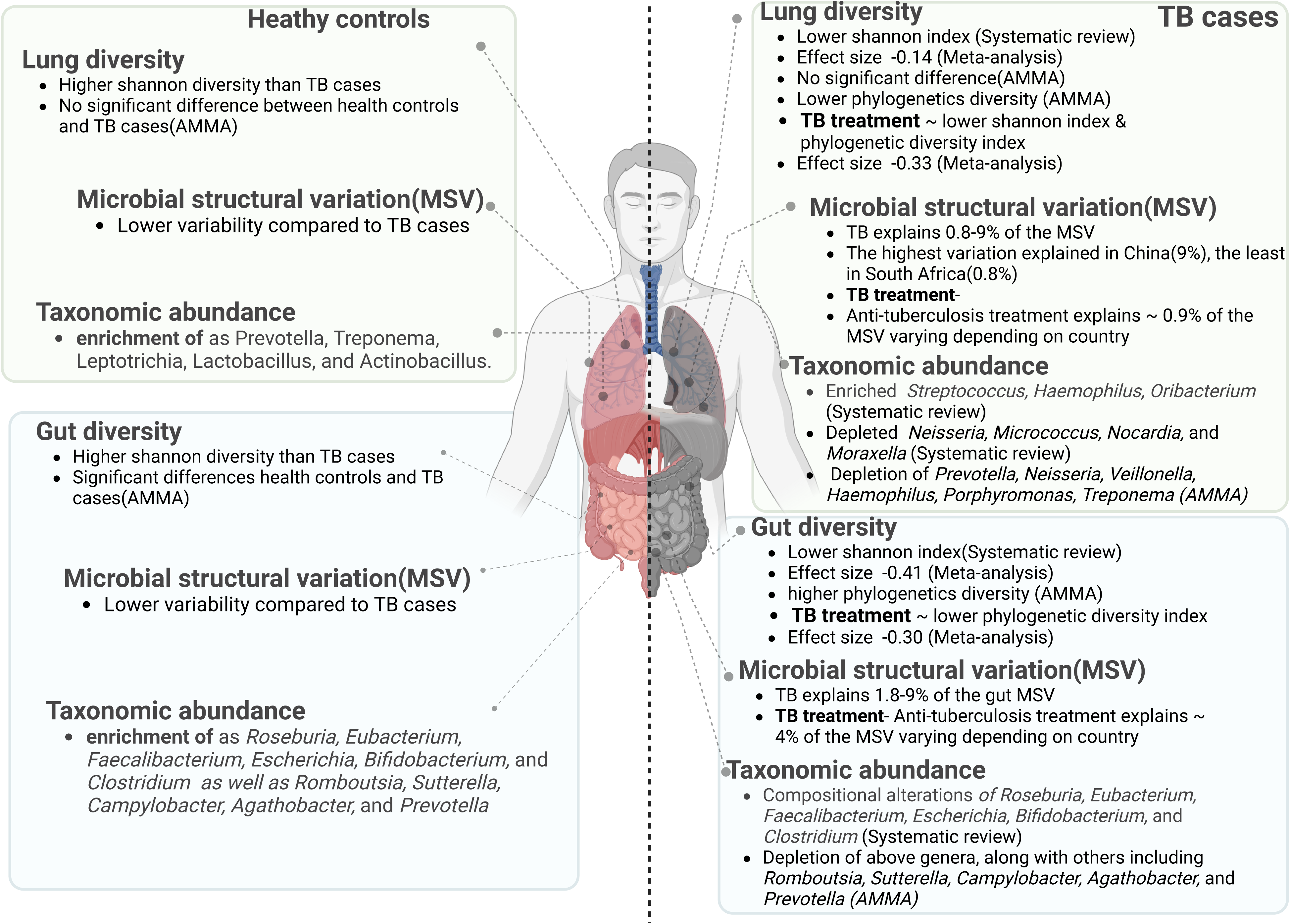
Summary of major findings and clinical relevance. This figure illustrates differences in lung and gut microbiota diversity, microbial structural variation, and taxonomic abundance between TB cases and healthy controls. It also highlights the impact of anti-TB treatment across multiple levels of microbiome analysis, providing a synthesized view of potential diagnostic and therapeutic implications.

## Conclusions

Our findings show that TB is generally associated with reduced microbial diversity in the lung, but not in the gut, with stronger associations observed within countries. In contrast, TB treatment leads to decreased diversity in both the lung and gut. These shifts are accompanied by taxonomic rewiring, which may have immunological, prognostic, and diagnostic relevance. Future research should aim to translate these insights into clinically actionable tools.

## Abbreviations

TB: Tuberculosis
Mtb: Mycobacterium tuberculosis
16S rRNA: 16S Ribosomal Ribonucleic Acid
PRISMA: Preferred Reporting Items for Systematic Reviews and Meta-Analyses AMMA: Amplicon-metagenomics meta-analysis
ASVs: Amplicon Sequence Variants
PCA: Principal Component Analysis
PERMANOVA: Permutational Multivariate Analysis of Variance PD: Faith’s Phylogenetic Diversity

## Declarations

### Ethics approval and consent to participate

Not applicable. This study is a systematic review and meta-analysis based entirely on data from previously published studies that are publicly available. No new data were collected from human participants, and no identifiable personal data were used. Therefore, ethical approval and informed consent were not required.

### Clinical trial

Clinical trial number: not applicable

## Consent for publication

Not applicable

## Availability of data and material

All data used in this study were obtained from publicly available sources. For the systematic review, data were extracted from published studies and summarized in the supplementary file: Supplementary_Data.xlsx. For the meta-analysis and amplicon-based metagenomic meta-analysis, we used 16S rRNA sequencing datasets retrieved from open-access repositories, including the NCBI Sequence Read Archive (SRA) and the European Nucleotide Archive (ENA). A complete list of accession numbers (study_accession_numbers.csv), along with combined sample-level metadata (*combined_metadata.tsv*), is available in the GitHub repository: https://github.com/MonicaMbabazi/16S_Analysis_Workflow (folder: *Global_Dataset.zip*).

## Authors’ contributions

1. MM, FN, JNW, and NM conducted the article search and screened studies to identify those eligible for inclusion. DPK contributed by resolving discrepancies that arose during the screening process.

2. MM and DPK developed the review protocol and submitted it for registration with PROSPERO.

3. MM, AM, BW, and WEJ performed the meta-analysis and metagenomic meta-analysis presented in this paper. MM, DPK, and AM prepared the first draft of the manuscript.

4. MO, IA, AA, and AM assisted in data interpretation and clinical relevance of the data.

5. All authors reviewed and contributed to the final version of the manuscript for submission.

## Data Availability

All data used in this study were obtained from publicly available sources. For the systematic review, data were extracted from published studies and summarized in the supplementary file: Supplementary_Data.xlsx. For the meta-analysis and amplicon-based metagenomic meta-analysis, we used 16S rRNA sequencing datasets retrieved from open-access repositories, including the NCBI Sequence Read Archive (SRA) and the European Nucleotide Archive (ENA). A complete list of accession numbers (study_accession_numbers.csv), along with combined sample-level metadata (combined_metadata.tsv), is available in the GitHub repository: https://github.com/MonicaMbabazi/16S_Analysis_Workflow (folder: Global_Dataset.zip).

## Acknowledgements

Not applicable

