## Supplemental Table 1 for "The impact of tuberculosis and its treatment on the lung and gut microbiota: A global systematic review, meta-analysis, and amplicon-based metagenomic meta-analysis"

**Table S1: MeSH terms**

| **Category** | **MeSH Terms/Keywords** |
| --- | --- |
| **Tberculosis-related** | ‘tuberculosis’ OR ‘mycobacterium tuberculosis’ OR ‘TB infection’ OR ‘TB disease’ |
| **Lung-Microbiome-related** | (‘lung’ OR ‘pulmonary’ OR ‘respiratory’ OR ‘sputum’) AND (‘microbiome’ OR ‘microbiota’ OR ‘microbial community’ OR ‘bacterial community’) |
| **Gut-Microbiome-related** | (‘gut’ OR ‘intestinal’ OR ‘stool’ OR ‘fecal’) AND ((‘microbiome’ OR ‘microbiota’ OR ‘microbial community’ OR ‘bacterial community’) |
