## Supplemental Table 2 for "The impact of tuberculosis and its treatment on the lung and gut microbiota: A global systematic review, meta-analysis, and amplicon-based metagenomic meta-analysis"

**Table S2: Summary of studies on the respiratory tract microbiota in pulmonary TB (TB cases Vs healthy individuals) ***

| **Study & Year** | **Country** | **Participants, Sample size, Sample type, Sequencing approach** | **Key findings** |
| --- | --- | --- | --- |
| Cui et al., 2012 | China | Participants: 31 TB patients & 24 healthy controls  Sample: Sputum & respiratory secretions of controls  Sequencing: *16S rRNA* (V3) | **Diversity (Shannon index):**   - ↑TB cases (6.19) - *↓*Healthy controls (5.51)   **Composition:**   - *TB cases:*   *Unique: Stenotrophomonas, Cupriavidus, Pseudomonas, Thermus, Sphingomonas, Methylobacterium,Diaphorobacter, Comamonas,* and *Mobilicoccus*.  *↑Anoxybacillus, Klebsiella, Acinetobacter, Pilibacter, Abiotrophia, Paucisalibacillus, and Rothia.*  *↓Neisseria, Porphyromonas, Parvimonas, Campylobacter, Haemophilus, and Fusobacterium.* |
| Wu, J et al., 2013 | China | Participants: 75 TB patients and 20 healthy controls  Sample: Sputum & throat swabs  Sequencing: *16S rRNA* (V1-V2) | **Diversity:**   - *↓*TB cases - ↑Healthy controls   ***Composition:***   - TB cases:   ↑*Firmicutes, Actinobacteria and Spirochaetes.*  ↑ *Streptococcus, Granulicatella* and *Pseudomonas.*  Unique: *Bergeyella and Sharpea.*   - **Controls:**   *↑Bacteroidetes and Fusobacteria.*  *↑Prevotella, Leptotrichia, Treponema, Catonella and Coprococcus* |
| Krishna et al., 2016 | India | Participants: 25 TB cases & 16 controls  Sample: Sputum  Sequencing: *16S rRNA* (V6-V7) | **Diversity (Shannon index):**   - *↓*TB cases (3.88) - *↑*Healthy controls (4.13)   ***Composition****:*   - TB cases:   ↑*Firmicutes and Actinobacteria*  ↑*Gammaproteobacteria, Streptococcus, Neisseria, Haemophilus,*  *Rothia, Leuconostoc.*  *↑Opportunistic pathogenic microbiota*   - Controls:   ↑*Proteobacteria and Fusobacteria.*  ↑*Actinobacillus* |
| Hu, Y., et al., 2020 | China | Participants: 93 TB patients & 70 controls  Sample: BAL  Sequencing: *16S rRNA* (V3-V4) | **Diversity (Shannon index):**   - *↓*TB cases (3.7) - *↑*Healthy controls (4.2)   ***Composition:***   - TB cases   ↑*Firmicutes.*  ↑*Mycobacteriaceae and Bacillaceae.*  ↑*Anoxybacillus/bacillus.*   - Controls   ↑*Proteobacteria and Bacteroidetes.*  ↑*Prevotellaceae, Veillonellaceae, and Bacillales.*  *↑Prevotella, Alloprevotella, Veillonella, and Gemella.* |
| Ding et al., 2021 | China | Participants: 123 TB patients, no healthy controls.  Sample: BAL  Sequencing:  *16S rRNA* & Shotgun | **Diversity (Shannon index):**   - ↓Bacteriologically confirmed TB patients (BC) - ↑Bacteriologically negative TB patients (BN)   ***Composition:***   - BC group   ↑*Actinobacteria*  ↑ *Prevotella, Streptococcus, and Haemophilus parainfluenzae*   - BN group   *↑Proteobacteria*  *↑Neisseria and Actinomyces* |
| Kateete et al., 2021 | Uganda | Participants: 192 TB patients, no healthy controls, 30 patients were followed during treatment.  Sample: Sputum  Sequencing:  *16S rRNA* (V3-V4) | **Diversity (Shannon index):**   - *↑*Untreated group (4.18) - ↓Treated group (3.65) - *↑*HIV-/TB cases - ↓HIV+/TB cases   ***Composition:***   - *Core TB microbiota: Streptococcus, Veillonella, Neisseria, Fusobacterium, Lachnoanaerobaculum, Atopobium, Peptostreptococcus and Leptotrichia.* |
| Lin, D et al., 2021 | China | Participants: 165 TB patients, no healthy controls.  Sample: Sputum  Sequencing:  *16S rRNA* (V4-V5) | **Diversity (Chao1):**   - ↓Drug Susceptible TB (DS) - *↑*Drug Resistant TB (DR)   ***Composition:***   - DR group:   *↑Proteobacteria*  *↑Ralstonia, Delftia, Campylobacter, and Neisseria*  ↓*Ralstonia, Prevotella, Alloprevotella, and Veillonella* |
| Valdez-Palomares et al., 2021 | USA | Participants: 39 TB patients & 6 healthy controls  Sample: Sputum  Sequencing:  *16S rRNA* (V3-V4) | **Diversity (Shannon index)**   - ↓TB cases (4.4) - *↑*Healthy controls (4.8)   ***Composition:***   - TB cases:   *↑*Streptococcus, Neisseria, Prevotella 7, Moraxella, and Veillonella  *↑   opportunistic pathogens*   - Controls:   *↑Bacteroidetes, Patescibacteria, and Spirochaetes,*  *Streptococcus*.  *↑Prevotella 7, Veillonella, Prevotella, and Alloprevotella* |
| Ticlla et al., 2021 | Tanzania | Participants: 334 TB patients & no  controls  Sample: Sputum  Sequencing:  *16S rRNA* (V3-V4) & shotgun | **Diversity (Faith’s PD)**   - *↑*TB patients with abnormal CXR - ↓TB patients with normal CXR   ***Composition:***   - Underweight patients:   ↑*Selenomonas and Fusobacterium*  ↓*Streptococcus*   - HIV+/TB cases:   *↑ herpesviruses* and *anelloviruses* |
| Ueckermann et al, 2022 | South Africa | Participants: 71 patients &   no healthy controls  Sample: Sputum  Sequencing:  *16S rRNA* (V3-V4) | **Diversity (Shannon index)**   - *↑*HIV-/TB - ↓HIV+/TB   ***Composition:***   - TB cases:   *↑Proteobacteria, Firmicutes, Actinobacteria, and Bacteroidetes*. |
| Cai et al, 2022 | China | Participants: 30 TB patients and 30 healthy controls  Sample: Sputum  Sequencing:  *16S rRNA* (V1-V3) | **Diversity (Shannon index)**   - ↓TB cases (2.63) - *↑*Healthy controls (3.08)   ***Composition:***   - TB cases:   *↑Firmicutes, Actinobacteria, Proteobacteria, Fusobacteria, TM7, and Spirochaetes*  *↑Rothia*   - Controls:   *↑Bacteroidetes, Patescibacteria, and Spirochaetes,*  *Streptococcus*.  *↑Streptococcus,* and *Veillonella* |
| Y.Huang et al, 2022 | China | Participants: 30 TB patients & 18 healthy controls  Sample: Nasal swab  Sequencing:  *16S rRNA* (V3-V4) | **Diversity (Shannon index)**   - *↑*TB cases (1.5) - ↓Healthy controls (1.2)   ***Composition:***   - TB cases:   *↑Proteobacteria*  *↑Pseudomonadales and Moraxellaceae*   - Controls:   *↑Bacillales, and Lachnospiraceae* |
| M.Zhang et al, 2022 | China | Participants: 37 TB patients, 14 TB patients were treated, and 13 healthy controls  Sample: BAL  Sequencing:  *16S rRNA* (V3-V4) | **Diversity (Shannon index)**   - *↑*TB cases not treated (10.2) - ↓Healthy controls (9.1) - ↓Treated group-1 Month (8.2)   ***Composition:***   - TB cases:   *↑Proteobacteria*  *↑Oscillospira, Streptococcus, and Allobaculum*   - Controls:   *↑Firmicutes*  *↑Prevotella,  Akkermansia, and Ruminococcus*   - *Anti-TB treatment group:*   *↑Bacteroidetes and Firmicutes*  ↓*Proteobacteria and Actinobacteria*  *↑Bacteroides, Ruminococcus, and Oscillospira* |
| Xiao et al, 2022 | China | Participants: 45 TB patients (12 untreated, 15 treated, 11 cured TB patients) & 8 healthy controls  Sample: BAL  Sequencing:  *Shotgun* | **Diversity (Shannon index)**   - ↓TB cases (2.1) - *↑*Healthy controls (2.5) - ↓Untreated TB group (2.1) - *↑*Treated TB group (2.5)   ***Composition:***   - *Healthy and cured individuals show no significant composition* |
| Musisi et al, 2023 | Tanzania | Participants: 397 TB patients and no healthy controls. 9 pre and 9 post-treated TB patients.  Sample: Sputum  Sequencing:  *16S rRNA* (V3-V4) | **Diversity (Shannon index):** HR35mg/kgZE   - *↑Untreated* TB group (5·49) - ↓*Treated* TB group, week2 (4·76)   ***Composition:***   - *Pre-treated* TB group:   *↑Firmicutes, Bacteroidetes, Proteobacteria, and Actinobacteria*  *↑Streptococcus*   - *Treated* TB group*:*   ↓Neisseria |
| Ruiz-Tagle et al, 2023 | Chile | Participants: 51 TB patients & 31 healthy controls  Sample: Nasal swab  Sequencing:  *16S rRNA* (V1-V3) | **Diversity (Shannon index)**   - ↓TB cases (2.64) - *↑*Healthy controls (3.23)   ***Composition:***   - TB cases:   *↑Reyranella, Polaromonas, Nocardioides, Micromonospora, Micrococcus, Deinococcus, and Acidovorax*   - Controls:   *Unique: Finegoldia, Romboutsia, Dolosigranulum, Mesrhizobium, Rickettsiella, Variovorax and Microbacterium*  *↑Reyranella and Micrococcus* |
| Liu et al., 2023 | China | Participants: 110 TB patients & 25 healthy controls  Sample: Lung tissue  Sequencing:  *Shotgun* | **Diversity (Shannon index)**   - *↑*TB cases (2.4) - ↓Healthy controls (1.8)   ***Composition:***   - TB cases:   *↑Pseudomonas pharmacofabricae, and Enterobius vermicularis*   - Controls:   *↑Staphylococcus epidermidis*, *S. aureus*, and *P. aeruginosa* |
| Cheung et al., 2013 | China | Participants: 22 TB patients & 14 healthy controls  Sample: Sputum  Sequencing:  *16S rRNA* (V1-V2) | **Diversity (Shannon index)**   - *similar in TB cases and controls*   ***Composition:***   - TB cases:   *↑Proteobacteria and Bacteroidetes*  *↑Mogibacterium, Moryella and Oribacterium*   - Controls:   *↑Firmicutes*  *↑Lactobacillales* |
| Sala et al 2020 | Multicenter: Switzerland, Italy, Bangladesh, Gambia, Vietnam, and Uganda | Participants:  15 TB patients & 15 healthy controls  Sample: Sputum  Sequencing:  *16S rRNA* (V1-V2) | **Diversity:**   - ↓TB cases (4.9) - *↑*Healthy controls (5.2)   **Composition:**   - TB cases   Most prevalent *Streptococcus and Prevotella*  *↑Neisseriaceae* |
| Botero et al., 2014 | Colombia | Participants: 6 TB patients & 6 healthy controls  Sample: Sputum & oropharynx  Sequencing:  *16S rRNA* (V1-V2) | **Diversity (Shannon index)**   - ↓TB cases (4.8) - *↑*Healthy controls (5.8)   ***Composition:***   - TB cases   *↑Streptococcaceae* |
| C.C. Naidoo et al, 2021 | South Africa | Participants: 105 TB patients & 155 healthy controls  Sample: Sputum & Stool  Sequencing:  *16S rRNA* (V4) | **Diversity (Shannon index)**   - ↓TB cases - *↑*Healthy controls   ***Composition:***   - TB cases:   *↑anaerobes (Paludibacter, Lautropia, Alloiococcus, Peptoniphilus)*  ↓*Alloscardovia* |
