## Supplemental Table 3 for "The impact of tuberculosis and its treatment on the lung and gut microbiota: A global systematic review, meta-analysis, and amplicon-based metagenomic meta-analysis"

**Table S3: A summary of studies on the gut microbiota in pulmonary TB**

| **Study** | **Country** | **Participants, Sample size, Sample type, Sequencing approach** | **Key findings** |
| --- | --- | --- | --- |
| A. Maji et al., 2017 | India | **Participants:** 6 TB patients & 6 healthy controls  **Sample:** Stool  **Sequencing:**  *16S rRNA* (V1) | **Diversity (Shannon index)**   - ***↑***TB cases (4.66) - ↓Healthy controls (3.79)   ***Composition:***   - **TB cases:**   ***↑***butyrate and propionate-producing bacteria like *Faecalibacterium*, *Roseburia*, *Eubacterium & Phascolarctobacterium*   - **Controls:**   ***↑****Prevotella* and *Bifidobacterium* |
| Luo et al., 2017 | China | **Participants:** 57 TB patients & 37 healthy controls  **Sample:** Stool  **Sequencing:**  *16S rRNA* (V4) | **Diversity (Shannon index)**   - ***↑***TB cases (5.7) - ↓Healthy controls (5.5)   ***Composition:***   - **TB cases:**   ***↑****Proteobacteria*  ↓*Bacteroidetes*  ↓*Prevotella*  *↑Escherichia* and *Streptococcus* |
| Hu et al 2019 | China | **Participants:** 46 TB patients & 31 healthy controls  **Sample:** Stool  **Sequencing:** Shotgun | **Diversity (Shannon index)**   - ↓TB cases (1.5) - ***↑***Healthy controls (2)   ***Composition:***   - **TB cases:**   ***↑****Coprobacillus* bacterium and *Clostridium bolteae*   - **Controls:**   ***↑***SCFA-producing bacteria (*Roseburia inulinivorans, R. hominis, R. intestinalis, Eubacterium rectale, and Bifidobacterium adolescentis*) |
| Namasivayam et al., 2020 | Mali | **Participants:** 21 MTB patients, 11 treated, & 10 healthy controls  **Sample:** Stool  **Sequencing:**  *16S rRNA* (V4) | **Diversity (Shannon index)**   - ***↑***Healthy controls (5.8) - ↓TB cases not treated (5.7) - ↓Treated group (4.4)   ***Composition:***   - **TB cases:**   ***↑****Enterobacteriaceae* |
| Wang J et al, 2020 | China | **Participants:** 58 TB patients (52 not treated and 6 treated), No healthy controls  **Sample:** Stool  **Sequencing:**  *16S rRNA* (V3-V4) | **Diversity (Shannon index)**   - ↓Untreated group (2.65) - ***↑***Treated group (2.3)   ***Composition:***   - **Treatment:**   induced a lasting gut microbiota dysbiosis |
| S. Wang et al., 2021 | China | **Participants:** 76 TB patients & no healthy controls  **Sample:** Stool  **Sequencing:**  *16S rRNA* (V3-V4) | **Diversity (Shannon index)**   - ↓TB cases (4.8) - ***↑***Healthy controls (5.2)   ***Composition:***   - **TB cases:**   ***↑****Bacteroides, Parabacteroides, Fusobacterium and Lachnoclostridium*   - **Controls:**   ***↑****Blautia*, *Roseburia*, *Bifidobacterium*, *Ruminococcaceae*, *Fusicatenibacter* and *Romboutsia* |
| Cao et al., 2021 | China | **Participants:** 29 TB patients & 22 healthy controls  **Sample:** Stool  **Sequencing:**  *16S rRNA* (V3-V4) | **Diversity (Simpson index, no Shannon diversity measure reported)**   - ↓TB cases - ***↑***Healthy controls   ***Composition:***   - **TB cases:**   ***↑****Anaerostipes*   - **Controls:**   *↑Eubacterium coprostanoligenes* and *Ruminococcus callidus* |
| Khaliq et al., 2021 | Pakistan | **Participants:** 42 TB patients &  40 healthy controls  **Sample:** Stool  **Sequencing:**  *16S rRNA* (V3-V4) | **Diversity (Shannon index)**   - ***↑***TB cases (3.6) - ↓Healthy controls (3.2)   ***Composition:***   - **TB cases:**   ***↑****Ruminococcaceae, Enterobacteriaceae, Erysipelotrichaceae, Bifidobacterium*   - **Controls:**   *↑*Prevotella |
| Shi W. et al., 2021 | China | **Participants:** 94 TB patients (55 untreated and 39 treated) & 62 healthy controls  **Sample:** Stool  **Sequencing:**  *16S rRNA* (V3-V4) | **Diversity (Shannon index)**   - ***↑***Healthy controls (5.0) - ↓TB cases and untreated (4.2) - ↓Treated group (3.8)   ***Composition:***   - **TB cases:**   ***↑****Enterococcus, Clostridioides, and Rothia.*   - **Controls:**   *↑Anaeroglobus, Lachnoclostridium, and Lachnospiraceae* |
| Yoon et al., 2022 | South Korea | **Participants:** 11 TB patients & 10 healthy controls  **Sample:** Stool  **Sequencing:**  *16S rRNA* (V3-V4) | **Diversity (Shannon index)**   - ***↑***Healthy controls - ↓TB cases - ↓Treated group   ***Composition:***   - **TB cases:**   ***↑****Verrucomicrobia.*  ***↑***Butyrate-producing bacteria (*Blautia* and *Roseburia*)  ↓*Proteobacteria* |
| Ding et al, 2022 | China | **Participants:** 10 TB patients & 20 healthy controls  **Sample:** Stool  **Sequencing:**  *16S rRNA* (V3-V4) | **Diversity (Shannon index)**   - ↓TB cases (2.4) - ***↑***Healthy controls (3.6)   ***Composition:***   - **TB cases:**   ***↑****Solobacterium and Actinobacteria*   - **Controls:**   ***↑****Prevotella, Romboutsia, Dialister, and Gemmiger*  ↓*[Granulicatella](https://www.sciencedirect.com/topics/pharmacology-toxicology-and-pharmaceutical-science/granulicatella), Solobacterium,  and* [*Erysipelotrichaceae*](https://www.sciencedirect.com/topics/immunology-and-microbiology/erysipelotrichaceae) |
| Wang et al, 2022 | China | **Participants:** 56 TB patients & 50 healthy controls  **Sample:** Stool  **Sequencing:**  *16S rRNA* (V3-V4) | **Diversity (Shannon index)**   - ↓TB cases (5.2) - ***↑***Healthy controls (6.9)   ***Composition:***   - **TB cases:**   ↓*Firmicutes and Tenericutes*   - **Controls:**   ***↑****Roseburia* |
| Ye et al, 2022 | China | **Participants:** 69 TB patients & 63 healthy controls  **Sample:** Stool  **Sequencing:**  *16S rRNA* (V4) | **Diversity (Shannon index)**   - ↓TB cases (4.6) - ***↑***Healthy controls (5.5)   ***Composition:***   - **TB cases:**   ***↑****Bacteroides*, *Parabacteroides*, and *Veillonella*  ↓*Faecalibacterium*, *Bifidobacterium*, *Agathobacter* and *CAG-352* |
| Luo et al, 2023 | China | **Participants:** 29 TB patients & 11 healthy controls  **Sample:** Stool  **Sequencing:**  *16S rRNA* (V3-V4) | **Diversity (Shannon index)**   - ↓TB cases (5.6) - ***↑***Healthy controls (6.7)   ***Composition:***   - **TB cases:**   ***↑****Faecalibacterium, Lachnoclostridium, Sellimonas, Streptococcus, Bacteroides, and R. gnavus*   - **Controls:**   ***↑****Enterococcus, R. torques, Parasutterella, Blautia, Subdoligranulum and Escherichia-Shigella* |
| Y.Huang et al, 2023 | China | **Participants:** 40 TB patients & 20 healthy controls  **Sample:** Stool  **Sequencing:**  *16S rRNA* (V3-V4) | **Diversity (Shannon index)**   - ↓TB cases (2.2) - ***↑***Healthy controls (3.2)   ***Composition:***   - **TB cases:**   ***↑****Faecalibacterium, Clostridia and Gammaproteobacteria*   - **Controls:**   *↑Bifidobacterium* and *Blautia* |
| Wipperman, et al., 2017 | USA | **Participants:** 44 TB patients (of these, 19 were untreated and 25 treated) & 50 healthy controls  **Sample:** Stool  **Sequencing:**  *16S rRNA* (V3-V4) | **Diversity (Shannon index)**   - ***↑***Healthy controls (3.41) - ↓TB cases not treated (3.34) - ↓Treated group (3.22)   ***Composition:***   - **TB cured individuals:**   *↓Bacteroidetes genera Bacteroides.*  ***↑****Faecalibacterium, Eubacterium, and Ruminococcus* |
| Wu et al., 2023 | China | **Participants:** 150 TB patients (60 on 6-month, 60 0n 12-month, and 30 no TB treatment) & no healthy controls  **Sample:** Stool  **Sequencing:**  *Shotgun* | **Diversity (Shannon index)**   - ↓Treated group (1.65) - ***↑***Not treated (1.80) |
| N.Seraphin et al, 2023 | USA | **Participants:** 6TB patients and they were followed during treatment, & 6 healthy controls  **Sample:** Stool  **Sequencing:**  *16S rRNA* (V1-V2) | **Diversity (Shannon index)**   - ↓TB cases (0.5) - ***↑***Healthy controls (0.6) - Treated group (0.2) - Untreated group (0.5) |
