## Supplementary figures and images for "The impact of tuberculosis and its treatment on the lung and gut microbiota: A global systematic review, meta-analysis, and amplicon-based metagenomic meta-analysis"

### Supplemental Fig 1

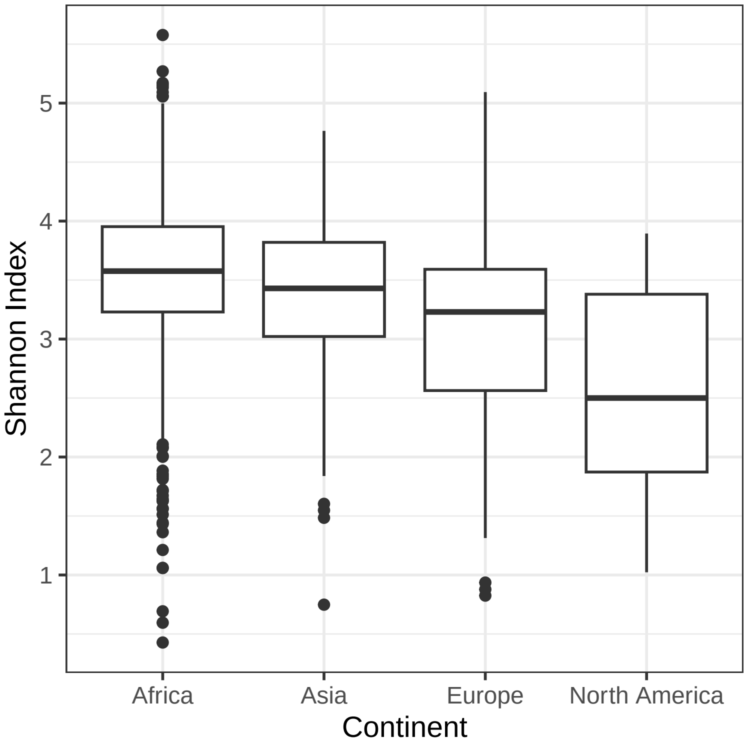
